# Plasma oxylipin profile discriminates ethnicities in subjects with Non-alcoholic steatohepatitis: An exploratory analysis

**DOI:** 10.1101/2021.12.01.21267151

**Authors:** Tagreed A. Mazi, Kamil Borkowski, Oliver Fiehn, Christopher L. Bowlus, Souvik Sarkar, Karen Matsukuma, Mohamed R. Ali, Dorothy A. Kieffer, Yu-Jui Y. Wan, Kimber L. Stanhope, Peter J. Havel, John W. Newman, Valentina Medici

## Abstract

Non-alcoholic fatty liver disease (NAFLD) includes a range of liver pathologies from steatosis (NAFL) to non-alcoholic steatohepatitis (NASH). With no clear mechanism, it affects Hispanics in the U.S. disproportionately compared to other ethnicities. Polyunsaturated fatty acids (PUFAs) metabolism and downstream inflammatory lipid mediators including oxylipin (OXL) and endocannabinoid (eCB) are altered in NAFLD and thought to contribute to its pathogenesis. It is not clear if variations in PUFA metabolism and downstream lipid mediators characterize ethnicity in NAFLD. This pilot study employed targeted lipidomic profiling for plasma PUFAs, non-esterified OXLs and eCBs in White Hispanics (HIS, *n* =10) and Caucasians (CAU, *n* =8) with obesity and biopsy-confirmed NAFL, compared with healthy control subjects (HC; *n* =14 HIS; n =8 CAU). Results indicate diminished long chain PUFA profile in HIS with NAFL and NASH, independent of obesity and histological severity. The profiling data also detected differences in plasma OXLs and eCBs profiles by ethnicity group in NASH, including lower levels of arachidonic acid derived OXLs observed in HIS. We conducted a secondary analysis to compare ethnicities within NASH (*n* =12 HIS; *n* =17 CAU). Results showed that plasma OXL profiles distinguished ethnicities with NASH and confirmed ethnicity-related differences in arachidonic acid metabolism. Our data also suggest lower lipoxygenase(s) and higher soluble epoxide hydrolase(s) activities in HIS compared to CAU with NASH. The underlying causes and implications of these differences on NAFLD severity are not clear and worth further investigation. Our findings provide preliminary data suggesting ethnicity-specific plasma lipidomic signature characterizing NASH that requires validation.

- There are differences in plasma PUFA and OXL seen in HIS, independent of obesity.
- In NAFL, HIS had lower plasma LC-PUFA, independent of histological severity.
- Compared to CAU, HIS with NASH had lower arachidonic acid derived OXLs.
- Compared to CAU, HIS with NASH showed lower LOX and up-regulated sEH pathway(s).
- Plasma OXL profiles can discriminate between HIS and CAU with NASH.

## 1. Introduction

Non-alcoholic fatty liver disease (NAFLD) is a chronic liver condition affecting one in four adults worldwide and this rate increases with coexisting components of metabolic syndrome [1]. Its histological presentation includes hepatocellular steatosis, or non-alcoholic fatty liver (NAFL) with a range of necroinflammation with or without fibrosis. When hepatocellular damage and ballooning are present with NAFL, this is clinically defined as non-alcoholic steatohepatitis (NASH) [2]. The pathogenesis of NAFLD is not fully elucidated. However, its onset involves an interplay between genetics and environmental factors with coexisting comorbidities while the progression to NASH appears to be provoked by multiple or parallel metabolic hits including oxidative stress and inflammation [3, 4]. In the U.S., the risk and severity of NAFLD vary among ethnic/racial groups with Hispanics (HIS) being affected disproportionately and presenting more frequently with advanced inflammation and fibrosis compared to other ethnicities [5–7]. The metabolic drivers underlying this disparity are not clear.

Polyunsaturated fatty acid (PUFAs) are bioactive lipids and precursors to inflammatory lipid mediators including oxylipins (OXLs) and endocannabinoids (eCBs). OXLs are produced from PUFAs by mono- and di-oxygenases, including lipoxygenases (e.g. 5-LOX, 12-LOXs, and 15-LOXs); cyclooxygenases (i.e. COX-1 and -2), and a variety of cytochrome P450s (CYPs) [8]. PUFAs can also undergo non-enzymatic oxygenation mediated by free radicals and the rate of this production is increased under oxidative stress [9]. In general, OXLs from n-3 PUFAs have anti-inflammatory or less pro-inflammatory effect compared to those derived from n-6 PUFAs [10, 11]. The fatty acid ethanolamide (i.e. N-acylethanolamides), one class of eCB, are synthesized by complex interactions of lipases and fatty acid amide hydrolase from PUFAs and membrane associated precursors [12]. Collectively, these lipids work through receptor-mediated mechanisms and likely contribute to NAFLD by modulating processes including lipogenesis, inflammation, and mitochondrial β-oxidation [8, 13].

Previous lipidomic analyses showed that NAFLD is associated with dysregulated PUFAs metabolism [14–18]. Alterations in circulating OXL and eCB profiles are reported in NAFLD and other liver pathologies. In fact, numerous lipid mediators have been shown to predict NAFL or NASH [16, 19–24]. However, metabolomic profiling in NAFLD with regards to ethnicity is limited. Our prior semi-quantitative lipidomic profiling study indicated ethnicity-specific differences in plasma PUFA profiles in subjects with NAFL, with higher abundance of linoleic and α-linolenic acid seen in Caucasians (CAU) compared to ethnicity-matched lean subjects [25]. In the same study, the progression to NASH was characterized by ethnicity-specific differences in hepatic lipidomic profiles with higher levels of saturated and unsaturated fatty acids seen in NASH-HIS. Ethnicity was not previously addressed in OXL and eCB profiling efforts. Examining such lipidomic differences between ethnicities may shed light on potential mechanisms modulating the disparity in NAFLD prevalence and severity.

The objective of this “proof-of-concept” study is to examine ethnicity-related changes in PUFAs and their downstream inflammatory mediators in a group of subjects with obesity and biopsy-confirmed NAFL and NASH. We employed targeted lipidomic analysis of plasma PUFAs, OXLs, and the N-acylethanolamides class of eCBs to compare HIS and CAU subjects with medically complicated obesity to ethnicity-matched lean healthy controls (HC). Profiles in subjects diagnosed with NASH were also compared to ethnicity and BMI-matched participants without NASH (0-NASH). In addition, we conducted a secondary analysis including prospectively collected subjects to compare OXL profile between ethnicities in NASH.

## 2. Subjects and methods

### 2.1. Subjects and samples

All subjects self-reported ethnicity as either HIS or CAU. HC subjects (*n* =22) were recruited via public posts. Plasma and liver samples form bariatric surgery patients with medically complicated obesity were retrieved from the biobank repository of the Division of Gastroenterology and Hepatology, UC Davis Medical Center. The primary cohort (*n* =18) consisted of subjects with NAFL and various degrees of necroinflammation. Only subjects with NASH were included in the secondary analysis (*n* =9) and this cohort was expanded with prospectively collected subjects diagnosed with NASH (*n* =20). Subject inclusion and exclusion criteria and details on data collection are described elsewhere [25]. Briefly, plasma samples were collected preoperatively, and liver tissue samples were collected by biopsy performed during bariatric surgery. Liver histopathological evaluations were performed in a blinded fashion in the UC Davis Medical Center Department of Pathology. Samples were scored according to the NASH Clinical Research Network (NASH-CRN) histology system, The NAFLD Activity Score (NAS) and fibrosis scores were calculated [26]. NASH diagnosis was determined using a diagnostic algorithm based on the semi-quantitative scoring of steatosis, inflammation, and fibrosis [27]. All subjects were consented and the Institutional Review Board at the University of California, Davis approved the study protocol (# 856052).

### 2.2. Targeted lipidomic analysis

Plasma quantitative lipidomic profiling of PUFAs, OXLs and NAEs was performed by ultra-high-performance liquid chromatography-electrospray ionization-tandem mass spectrometry (UPLC-ESI-MS/MS), as previously described [28]. Briefly, plasma samples were enriched with deuterated surrogates, isolated by liquid/liquid extraction, and separated and quantified by UPLC-ESI-MS/MS. In the primary analysis, ESI-polarity switching facilitated the simultaneous detection of eCBs (positive mode) and oxylipins and PUFAs (negative mode) on an API 6500 QTRAP (AB Sciex, Framingham, MA, USA). Metabolites were quantified against authentic analytical standards with 6-to-10-point calibration curves and calculated concentrations were corrected for analytical surrogate recovery. This method detected 5 PUFAs and 66 lipid mediators, including 10 eCBs and 46 OXLs.

The secondary analysis was done on an API 4000 QTrap (AB Sciex, Framingham, MA, USA) and restricted to the negative mode electrospray ionization to increase the power of the OXLs discovery. This approach detected 46 OXLs and 2 nitrolipids. Details on the analysis protocols and reported data are available on the Metabolomics Workbench (http://www.metabolomicsworkbench.org), ID numbers (ST000977 and ST001845). Analyses were carried out at the UC Davis West Coast Metabolomics Center. In this manuscript, abbreviations used for OXLs and eCBs follow standard consensus and are detailed with lipid identifiers in Table S1.

### 2.3. The ratio of n-6 to n-3-PUFAs and estimation of enzymatic surrogates

The ratio of n-6 ton-3 PUFAs was calculated as arachidonic (20:4n6)/eicosapentaenoic (20:5n3) + docosahexaenoic acids (22:6n3). The production rate of docosahexaenoic acid was estimated as docosahexaenoic/ (α-linolenic (18:3n3) + eicosapentaenoic acids. The product to substrate ratio was used to estimate multiple enzymatic indices. Fatty acid desaturase-1 (*FADS1*, Δ-5 desaturase) was estimated as arachidonic /dihomo-γ-linolenic acids (20:3n6). Various dihydroxy/epoxy regioisomeric ratios were calculated to provide soluble epoxide hydrolase (sEH) activity indices: 12,13-DiHOME/12(13)-EpOME; 9,10-DiHOME/9(10)-EpOME; 15,16-DiHODE/15(16)-EpODE; 12,13-DiHODE/12(13)-EpODE; 9,10- DiHODE/9(10)-EpODE; 17,18-DiHETE/17(18)-EpETE; and 14,15-DiHETrE/14(15)-DiHETrE.

### 2.4. Statistical analysis

Statistical analyses were performed using JMP Pro 14.1 (SAS Institute Inc., Cary, NC; http://www.jmp.com). Outliers were identified and excluded using “robust Huber M test”. Lipids with >30% missing data were excluded. Missing data were imputed by “multivariate normal imputation”. Data normality was achieved through application of Johnson’s transformation. To inspect for outliers, especially in subjects with stage 4 fibrosis (*n* =2), principal component analysis (PCA) was used to visualize data after processing and no outliers were detected (Figure S1).

Non-normalized data were used to calculate metabolite geometric means. Fold change (FC) was calculated for each ethnicity separately as (A - B)/B where A is the geometric mean of (NAFL or NASH) and B is the geometric mean of (HC or 0-NASH). A FC >0 indicates an increase and <0 indicates decrease and ± 20% FC was set as a threshold. Student’s t-test of Johnson normalized data was used to examine differences between (NAFL vs. HC) and (0-NASH vs. NASH) in each ethnicity. Full factorial analysis of covariates (ANCOVA) was employed to evaluate the interaction of ethnicity x health status. This model included ethnicity (HIS or CAU), health status (NAFL or HC; 0-NASH or NASH), ethnicity x health status interaction as fixed effects, with age and sex as covariates. To check any effect of fibrosis or advance NAS score on the differences observed, we repeated the analysis on a subset of histology-matched subjects (*n* =5 HIS and n =5 CAU). Pathways/network visualization with fold change and *p*-values were plotted using Cytoscape 3.8.2 (https://cytoscape.org) [29]. Mean differences were considered likely at *p* <0.05. To adjust for false discovery rate (FDR), Benjamini-Hochberg FDR correction was performed [30]. A *q* =0.2 was set as a threshold, given the pilot nature and small sample size of the study.

For the secondary analysis, raw data were auto-scaled to correct for batch effect (Figure S2) [31]. Lipids affected by batch were excluded (3 lipids) and data were normalized by Johnson’s transformation. Partial least square-discriminant analysis (PLS-DA) was performed to discriminate ethnicities in NASH subjects with leave- one-out cross validation (LOOCV) [32]. An R^2^ and Q^2^ > 0.5 are acceptable values to indicate reliability of the model in explaining differences between groups [33]. A variable importance in projection (VIP) score of >1.0 was set as a threshold for variable selection. To check any effect of advanced fibrosis, we repeated the analysis after excluding subjects with fibrosis grade 3 and 4 (*n* =3 HIS and n =4 CAU).

## 3. Results

### 3.1. Subject characteristics

The clinical and histological features of NAFL subjects from the primary cohort are presented in Table 1. The mean age was 47 ± 15 and 43 ± 14.2 in NAFL-HIS and HC-HIS groups, respectively; 50 ± 18 and 44 ± 12 in NAFL-CAU and HC-CAU, respectively (n.s). In HIS, the mean BMI was 46 ± 6 in NAFL and 26 ± 2.4 in HC (*p-*value <0.05). In CAU, the mean BMI was 42 ± 8 in NAFL and 25 ± 3 in HC (*p-*value <0.05). Within NAFL group, the mean NAS score was 3.2 ± 2.6 and 2.8 ± 1.4 for HIS and CAU, respectively (n.s). No difference in clinical and histological parameters was found between ethnicities. In subjects with NASH compared to 0-NASH, the mean NAS score was 4.8 ± 1.7 and 4 ± 1 for HIS and CAU, respectively (n.s) (data not shown).

**Table 1.** Demographic, clinical, and histological characteristics of study subjects in primary analysis. General characteristics of NAFL and HC groups in both ethnicities shown as percent (for categorical data) and mean ± SD (for nominal data). Comparisons were performed by *t*-test (nominal) or chi-square test (categorical). ALT, alanine aminotransferase; AST, aspartate aminotransferase; BMI, body mass index; DM, diabetes mellitus; FBG, fasting blood glucose; HbA1c, hemoglobin A1c; HDL, high-density lipoprotein, low density lipoprotein; NAS, the NAFLD Activity Score; TG, triglycerides. *^(a)^* NAFL-HIS vs. HC-HIS; *^(b)^* NAFL-CAU vs. HC-CAU; *^(c)^* NAFL-HIS vs. NAFL-CAU.

|  | NAFL-HIS | NAFL-CAU | *p*-value * |
| --- | --- | --- | --- |
| Plasma, *n* (F/M) | 10 (7/3) | 8 (4/4) |  |
| Age (Yrs.) | 47 ± 15 | 50 ± 18 | 0.6 |
| DM, yes (%) | 4 (40) | 4 (50.0) | 1.0 |
| FBG mmol/L | 101 ± 14 | 94 ± 13 | 0.3 |
| Cholesterol (mg/dL) | 172 ± 28 | 166 ± 34 | 0.7 |
| TG (mg/dL) | 135 ± 79 | 129 ± 70 | 0.8 |
| HDL (mg/dL) | 44 ± 6 | 43 ± 8 | 0.7 |
| LDL (mg/dL) | 107 ± 24 | 99 ± 35 | 0.4 |
| HbA1c (%) | 6 ± 1 | 6 ± 1 | 0.7 |
| AST (U/L) | 31 ± 13 | 26 ± 9 | 0.6 |
| ALT(U/L) | 40 ± 27 | 31 ± 11 | 0.9 |
| Platelet | 280 ± 84 | 302 ± 110 | 0.8 |
| Steatosis | 23 ± 25 | 20 ± 15 | 0.9 |
| NAS | 3.2 ± 2.62 | 2.75 ± 1.41 | 0.7 |
| Steatosis (%) |  |  |  |
| < 5% | 4 (40) | 1(12.5) | 0.9 |
| 5 to ≤ 33% | 3 (30) | 5(62.5) |  |
| 34 to ≤ 66 % | 2 (20) | 2 (25.0) |  |
| > 66% | 1 (10.0) | 0 (0.0) |  |
| Inflammation (%) |  |  |  |
| None | 3 (30) | 1 (12.5) | 0.6 |
| Mild | 1 (10) | 4 (50.0) |  |
| Moderate | 4 (40) | 3 (37.5) |  |
| Severe | 2 (20) | 0 (0.0) |  |
| Ballooning (%) |  |  |  |
| None | 4 (40) | 5 (62.0) | 0.3 |
| Few | 5 (50) | 3 (37.5) |  |
| Many | 1 (10) | 0 (0.0) |  |
| Fibrosis (%) |  |  |  |
| None | 7 (70) | 7 (87.5) | 0.3 |
| 1A | 0 (0.0) | 1 (12.1) |  |
| 2 | 1 (10.0) | 0 (0.0) |  |
| 4 | 2 (20.0) | 0 (0.0) |  |

The secondary cohort included NASH subject with various degrees of necroinflammation and fibrosis (Table 2). When comparing NASH-HIS and NASH-CAU, no difference was found with BMI and other clinical and histological parameters. The mean NAS score was 5.1 ± 2 and 5.1 ± 1.3 for NASH-HIS and NASH-CAU, respectively (n.s).

**Table 2.** Demographic, clinical, and histological characteristics of NASH subjects in secondary analysis. General characteristics of subjects included in the secondary analysis shown as percent (for categorical data) and mean ± SEM (for nominal data). Comparisons were performed by *t*-test (nominal) or chi-square test (categorical). BMI - body mass index; CAU - Caucasian; HIS - Hispanic; NASH - Non-alcoholic steatohepatitis; NAS - the NAFLD Activity Score. (*) NAFL-HIS vs. NAFL-CAU.

|  | NASH-HIS |  |  | NASH-CAU |  |  | *p*-value |
| --- | --- | --- | --- | --- | --- | --- | --- |
| Plasma, *n* (F/M) | 12 (9/3) |  |  | 17 (11/6) |  |  |  |
| Age (Yrs.) | 49.4 | ± | 8.1 | 50.4 | ± | 13.3 | 0.816 |
| BMI (Kg/m2) | 41 | ± | 7.65 | 36.6 | ± | 7.35 | 0.585 |
| NAS | 5.08 | ± | 1.98 | 5.06 | ± | 1.3 | 0.97 |
| Steatosis (%) |  |  |  |  |  |  |  |
| < 5% | 2(16.7) |  |  | 0(0) |  |  | 0.363 |
| 5 to ≤ 33% | 4(33.3) |  |  | 7(41.2) |  |  |  |
| 34 to ≤ 66 % | 3(25) |  |  | 4(23.5) |  |  |  |
| > 66% | 3(25) |  |  | 6(35.3) |  |  |  |
| Inflammation (%) |  |  |  |  |  |  |  |
| None | 0 |  |  | 0(0) |  |  | 0.147 |
| Mild | 2(16.7) |  |  | 3(17.7) |  |  |  |
| Moderate | 6(50.0) |  |  | 13(76.5) |  |  |  |
| Severe | 4(33.3) |  |  | 1(5.8) |  |  |  |
| Ballooning (%) |  |  |  |  |  |  |  |
| None | 0(0) |  |  | 0(0) |  |  | 0.56 |
| Few | 8(66.7) |  |  | 13(76.5) |  |  |  |
| Many | 4(33.3) |  |  | 4(23.5) |  |  |  |
| Fibrosis (%) |  |  |  |  |  |  |  |
| None | 5(41.6) |  |  | 4(23.4) |  |  | 0.766 |
| 1a, b, c | 2(16.8) |  |  | 6(35.3) |  |  |  |
| 2 | 2(16.8) |  |  | 3(17.7) |  |  |  |
| 3 | 2(16.8) |  |  | 2(11.8) |  |  |  |
| 4 | 1(8.3) |  |  | 2(11.8) |  |  |  |

### 3.2. Ethnicity-related alterations in plasma PUFAs and lipid mediator profiles characterize NAFL

We examined differences in plasma fatty acids and lipid mediators between NAFL and HC (Figure 1 and Table S2). Compared to corresponding HC, 25 (38% of total detected) and 7 (11%) lipid levels were found different in NAFL-HIS and NAFL-CAU, respectively (FDR-adjusted *p* <0.2). Ethnicity-specific changes observed in NAFL, with interaction ethnicity x NAFL, include 8 lipids (15%) and 2 enzymatic ratios (raw *p-*Interaction <0.05) but did not survive FDR-correction (*q* =0.2). As NAFL-HIS presented with more NASH and fibrosis, and to rule out any effect of histological severity on the differences observed between ethnicities, the analysis was repeated on a subset of histology-matched subjects (Table S3). As a result, 12 lipids (19%) and one enzymatic ratio were found altered (raw *p-*Interaction <0.05), with 3 lipids (i.e., α-linolenic, linoleic acids, and 9-HpODE) surviving FDR correction.

**Figure 1.**
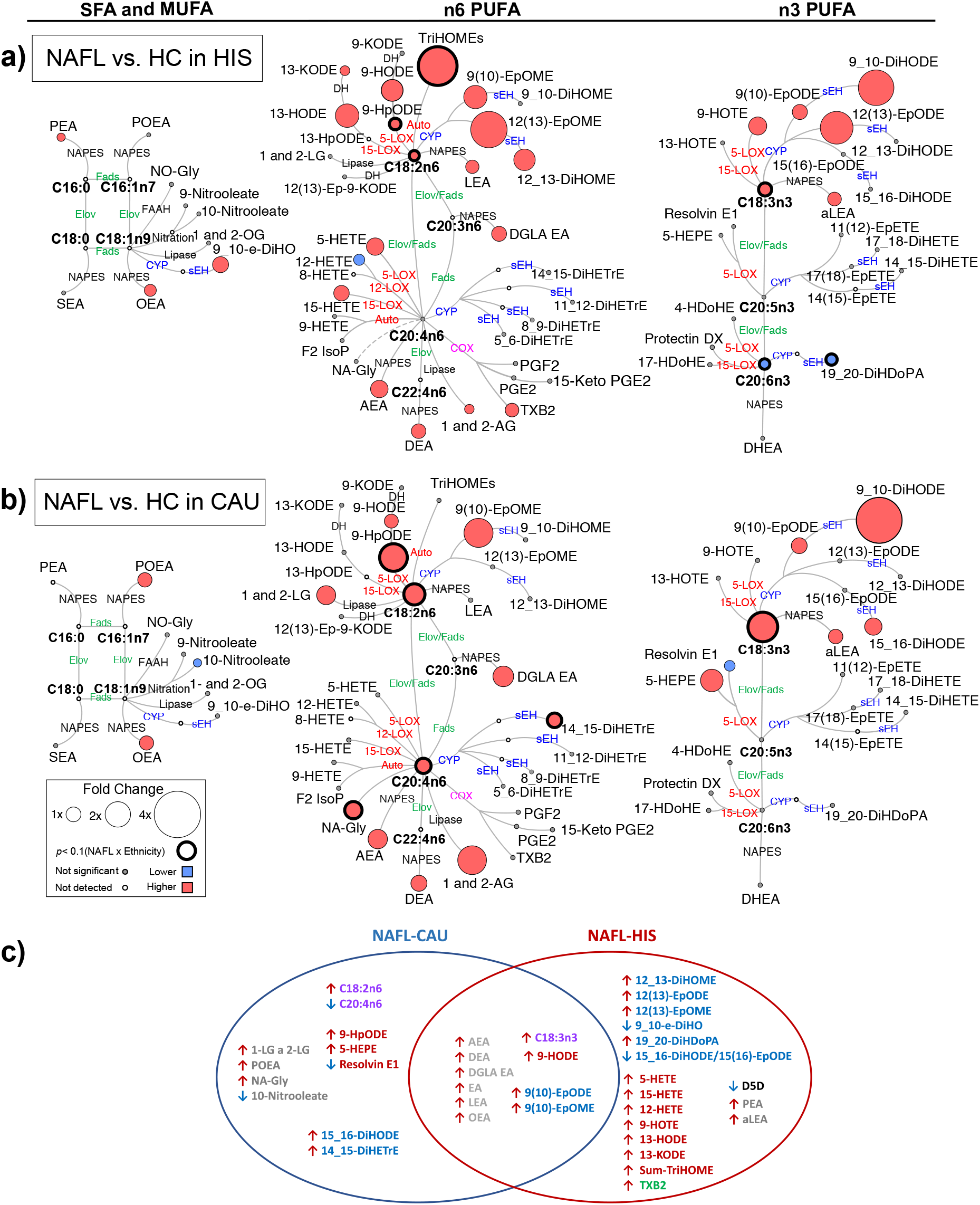
Differences in plasma PUFA and lipid mediators between NAFL compared to HC in primary cohort. Metabolic network for (a) HIS; (b) CAU illustrating saturated (SFAs), monounsaturated (MUFAs) and polyunsaturated fatty acids (PUFAs), including n-3 and n-6 fatty acids with pathway of oxylipins and endocannabinoids synthesis. Node size represents the fold change of lipids, calculated for each ethnicity separately as (HC - NAFL)/HC. Node’s color represents the directionality of metabolite differences: higher in NAFL (red); lower in NAFL (blue); no change (grey). Only fold change for lipids with differences between NAFL vs. HC (*t*-test raw *p* <0.05) and/or with interaction (ethnicity x NAFL) (ANCOVA raw *p* <0.05) are shown. Lipids with interaction (ethnicity x NAFL) (ANCOVA raw *p* <0.05) are marked with a solid circle and lipids. Lipids means and corresponding *p*-values are detailed in Table S3. NAFL (*n* =10 HIS and 8 CAU); HC (*n* =14 HIS and 8 CAU). (b) Venn-Diagram illustrating overlapping and unshared differences in plasma PUFA and lipid mediators between NAFL and HC in both ethnicities. Metabolites with FDR-adjusted *p* <0.2 are shown. Fatty acids are shown in (purple). Other metabolites are colored according to their metabolizing enzymes pathways, lipoxygenase, and autoxidation (red); cytochrome p450 epoxygenase and soluble epoxide hydrolase (blue); cyclooxygenase (green); N- acylphosphatidylethanolamide-phospholipase D (grey); other (black). Fatty acids are described by number of carbons and double bounds of the fatty acyl moiety (i.e., C18:2n6). CAU, Caucasian; HIS, Hispanics; HC, Healthy control; NAFL, Non-alcoholic fatty liver; ADH, alcohol dehydrogenase; AEA, arachidonoyl ethanolamine; AG, arachidonoyl glycerol; Auto, autoxidation; COX, cyclooxygenase; CYP, cytochrome P450; DEA, docosatetraenyl ethanolamide; DGLEA, dihomo-gamma-linolenoyl ethanolamide; DH, dehydrogenase; DHEA, docosahexaenoyl ethanolamide; DiHDoPA, dihydroxydocosapentaenoic acid; DiHETE, dihydroxyeicosatetraenoic acid; DiHETrE, dihydroxyeicosatrienoic acid; DiHO, dihydroxyoctadecanoic acid; DiHODE, dihydroxyoctadecadienoic acid; DiHOME, dihydroxyoctadecenoic acid; Elov, fatty acid elongase; Ep-KODE, epoxyoxooctadecenoic acid; EpETE, epoxyeicosatetraenoic acid; EpODE, epoxyoctadecadienoic acid; EpOME, epoxyoctadecenoic acid; F2-IsoP, F2 isoprostanes; FAAH, fatty acid amide hydrolase; Fads, fattyaciddesaturase; HDoHE, hydroxydocosahexaenoic acid; HEPE, hydroxyeicosapentaenoic acid; HETE, hydroxyeicosatetraenoic acid; HODE, hydroxyoctadecadienoic acid; HOTE, hydroxyoctadecatrienoic acid; HpODE, hydroperoxyoctadecadienoic acid; KETE, keto-eicosatetraenoic; KODE, keto-octadecadienoic acid; LEA, Linoleyl ethanolamine; LG, linoleoylglycerol; LOX, lipoxygenase; MUFA, Monounsaturated fatty acid; NA-Gly, arachidonylglycine; NAPES, N-acylphosphatidyl ethanolamine- specific; NO-Gly; OEA, oleoyl ethanolamine; OG, oleoylglycerol; PEA, palmitoyl ethanolamine; PGE, prostaglandin E; PGF, prostaglandin F; POEA, palmitoleoyl ethanolamide; PUFA, Polyunsaturated fatty acids; SEA, stearoyl ethanolamide; sEH, soluble epoxide hydrolase; SFA, Saturated fatty acids; TriHOME, trihydroxyoctadecaenoic acid; TXB, thromboxane.

When compared to HC, there were overlapping differences seen in NAFL for both ethnicities as well as ethnicity-specific differences (Figure 1). In both ethnicities, NAFL showed higher levels of several eCBs derived from PUFA and other fatty acids. The 18 carbon (C18) PUFAs, α-linolenic and linoleic acids showed similar higher trend with higher levels of downstream fatty acid alcohols, hydroperoxide, ketones, epoxides, and vicinal diols. Specific to NAFL-HIS, there were differentially higher levels of linoleic acid-triols (i.e., TriHOMEs; raw *p-*Interaction <0.05). On histology-matched analysis, a higher TriHOMEs and linoleic acid epoxide, 12(13)-EpOME, were found significant (raw *p-*Interaction <0.05). Of note, the n-6 to n-3 ratio was higher, however, with no ethnicity x NAFL interaction. In NAFL-CAU, there was differentially higher α-linolenic and linoleic acid and its hydroperoxide, 9-HpODE (raw *p-*Interaction <0.05). On histology adjustment analysis these lipids retained significance with linoleic acid, and 9-HpODE passing the FDR-threshold.

The 20 carbon (C20) and longer chain PUFAs (LC-PUFA), showed opposite trends with higher levels in NAFL-CAU and lower in NAFL-HIS. The ratio of docosahexaenoic/ eicosapentanoic + α-linolenic acids was found lower in both ethnicities (FDR-adjusted *p*-value). In HIS, arachidonic acid-derived alcohols, ketones, thromboxane were higher, however, with no interaction (ethnicity x NAFL). Docosahexaenoic acid and its vicinal diol, 19,20-DiHDoPA, levels were differentially lower (raw *p-*Interaction <0.05) with tendency shown for lower eicosapentanoic acid. On histology-matched analysis, lower levels of these lipids were found significant (raw *p-*Interaction <0.05). Specific to CAU, there was differentially higher arachidonic acid and its vicinal diol, 14,15-DiHETrE that remained significant after histology adjustment (raw *p* <0.05). While these findings show common alterations seen in NAFL for both ethnicities, they also highlight ethnicity-specific changes. This includes a divergence in LC-PUFA profile, mainly with lower eicosapentanoic and docosahexaenoic acids seen in HIS. Although these differences did not pass FDR adjustment, histology-matched analysis yielded consistent and stronger differences, suggesting ethnicity-specific differences characterized NAFL, independently of liver histology severity. It also suggests that fibrosis may weaken the differences between ethnicities.

### 3.3. Ethnicity-related differences in plasma PUFAs and lipid mediators’ independent of obesity

Ethnicity-specific differences in plasma fatty acids and lipid mediators within lean HC were examined (Figure S3 and Table S2). Among the differences observed, HIS had higher linoleic acid (raw *p*<0.05), α-linolenic acid, 9-HpODE, and TriHOMEs levels (FDR-adjusted *p*-value), and lower arachidonic acid-derived prostaglandin, PGE_2_ (FDR-adjusted *p*-value) levels. These findings indicate alterations in plasma PUFAs and lipid mediator profiles in HIS independent of obesity.

### 3.4. The progression to NASH is characterized by ethnicity-related alterations in plasma PUFAs and lipid mediator profiles

We examined differences in plasma fatty acids and lipid mediators between 0-NASH and NASH (Figure 2 and Table S4). Compared to corresponding 0-NASH, 7 (11% of total detected) and 6 (9%) lipids were found different in NASH-HIS and NASH-CAU, respectively (raw *p*-value <0.05). None passed the FDR-correction threshold. There were differentially altered lipids by ethnicity group (raw *p-*Interaction <0.05), including 11 lipids (17%) and two enzymatic ratios. Three of these lipids and the two enzymatic ratios passed the FDR-correction threshold.

**Figure 2.**
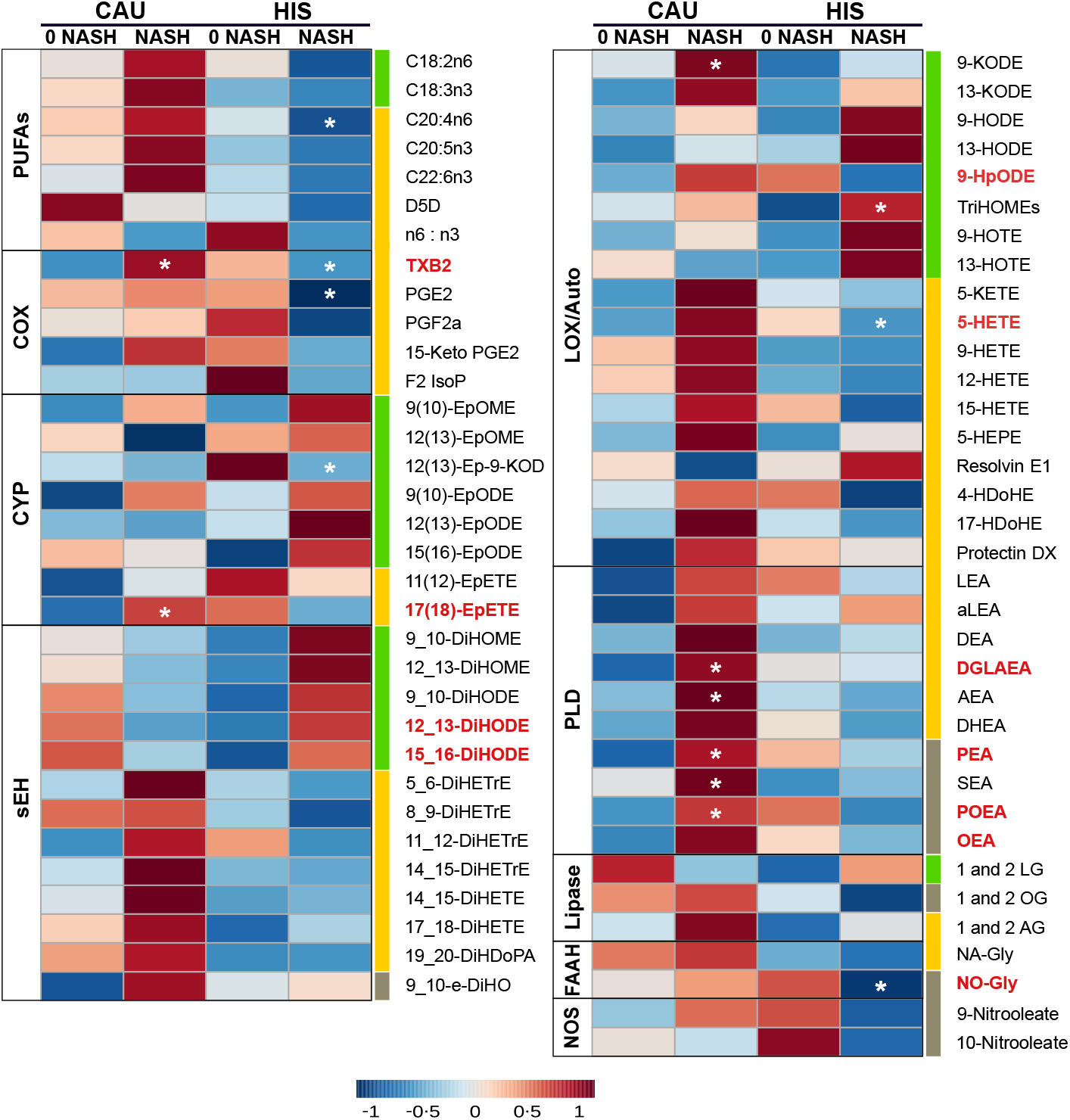
Heatmap illustrating fold changes and differences in plasma PUFA and lipid mediators between NASH compared to 0-NASH in primary cohort. Fold changes are indicated by color and intensity, with red indicating an increase, and blue indicating a decrease. Lipids different in NASH vs. 0_NASH (*t*-test raw *p* <0.05) are noted by (*). Lipids with ethnicity-related changes, or interaction (ethnicity x NASH), (ANCOVA raw *p* <0.05) are marked with bold red color. Fatty acids are described by number of carbons and double bounds of the fatty acyl moiety (i.e., C18:2n6). C18 PUFAs and related lipids are marked with “green”; LC-PUFAs and related lipids are marked with “yellow”; SFA and MUFA related lipids are marked with “grey”. Details on Lipids means, and corresponding *p*-values are detailed in Table S5. NASH (*n* =6 HIS and 3 CAU); 0-NASH (*n* =4 HIS and 5 CAU). Fatty acids are described by number of carbons and double bounds of the fatty acyl moiety (i.e., C18:2n6). CAU, Caucasian; HIS, Hispanics; NASH, Non-alcoholic steatohepatitis; ADH, alcohol dehydrogenase; AEA, arachidonoyl ethanolamine; AG, arachidonoyl glycerol; Auto, autoxidation; COX, cyclooxygenase; CYP, cytochrome P450; DEA, docosatetraenyl ethanolamide; DGLEA, dihomo-gamma-linolenoyl ethanolamide; DH, dehydrogenase; DHEA, docosahexaenoyl ethanolamide; DiHDoPA, dihydroxydocosapentaenoic acid; DiHETE, dihydroxyeicosatetraenoic acid; DiHETrE, dihydroxyeicosatrienoic acid; DiHO, dihydroxyoctadecanoic acid; DiHODE, dihydroxyoctadecadienoic acid; DiHOME, dihydroxyoctadecenoic acid; Elov, fatty acid elongase; Ep- KODE, epoxyoxooctadecenoic acid; EpETE, epoxyeicosatetraenoic acid; EpODE, epoxyoctadecadienoic acid; EpOME, epoxyoctadecenoic acid; F2-IsoP, F2 isoprostanes; FAAH, fatty acid amide hydrolase; Fads, fattyaciddesaturase; HDoHE, hydroxydocosahexaenoic acid; HEPE, hydroxyeicosapentaenoic acid; HETE, hydroxyeicosatetraenoic acid; HODE, hydroxyoctadecadienoic acid; HOTE, hydroxyoctadecatrienoic acid; HpODE, hydroperoxyoctadecadienoic acid; KETE, keto-eicosatetraenoic; KODE, keto-octadecadienoic acid; LEA, Linoleyl ethanolamine; LG, linoleoylglycerol; LOX, lipoxygenase; MUFA, Monounsaturated fatty acid; NA-Gly, arachidonylglycine; NAPES, N-acylphosphatidyl ethanolamine-specific; NOS - Nitric oxide synthase; NO-Gly; OEA, oleoyl ethanolamine; OG, oleoylglycerol; PEA, palmitoyl ethanolamine; PGE, prostaglandin E; PGF, prostaglandin F; PLD- N-acylphosphatidylethanolamide-phospholipase D; POEA, palmitoleoyl ethanolamide; PUFA, Polyunsaturated fatty acids; SEA, stearoyl ethanolamide; sEH, soluble epoxide hydrolase; SFA, Saturated fatty acids; TriHOME, trihydroxyoctadecaenoic acid; TXB, thromboxane.

With the progression from 0-NASH to NASH, less marked differences in plasma PUFA profile were observed. Compared to 0-NASH, NASH-HIS showed a trend for lower plasma PUFAs, only affecting arachidonic acid (raw *p*-value <0.05). There was a general trend for higher C18-PUFA derived alcohols, triol, epoxides and vicinal diols, with TriHOMEs being differentially higher (raw *p-*Interaction <0.05). We also observed a trend for lower LC-PUFA derived lipid mediators, mainly affecting arachidonic acid alcohols, 5-HETE; thromboxane, TXB_2_; and prostaglandin, PGE_2_. The levels of 5-HETE and TXB_2_ were found differentially changed between ethnicities in NASH (raw *p-*Interaction <0.05). Also, the oleic acid-derived N-oleoyl glycine levels were lower with NASH (raw *p-*Interaction <0.05).

In NASH-CAU, there was a trend of higher C18, LC-PUFAs and downstream lipid mediators. Interaction (ethnicity x NASH) was shown with higher 9-HpODE, TXB_2_, and in eicosapentaenoic acid epoxide, 17(18)-EpETE (raw *p-*Interaction <0.05). There was an opposite trend for vicinal diols derived from C18-PUFA that were higher in HIS and lower in CAU, compared to corresponding 0-NASH, with interaction (ethnicity x NASH) shown for 12,13- and 15,16-DiHODE (raw *p-*Interaction <0.05). Multiple sEH enzymatic indices were higher in HIS and lower in CAU, including 9_10-DiHOME/9(10)-EpOME and 9_10-DiHODE/9(10)-EpODE (raw *p-*Interaction <0.05). Also, with NASH, there were higher levels of many eCBs, including dihomo-γ-linolenoylethanolamide, palmitoleoylethanolamide, palmitoleoylethanolamide, oleoylethanolamide and N-oleoyl glycine (raw *p-*Interaction <0.05). Although PUFA changes are less marked in NASH, trends are consistent with changes seen in NAFL and support divergence in LC-PUFA profiles. It also highlights ethnicity-related differences in OXLs and eCBs associated with NASH progression. Given that NASH groups in both ethnicities had comparable NAS scores, this suggests the ethnicity-related differences observed with NASH are not likely driven by histological severity.

### 3.5. Plasma OXLs profile discriminate between ethnicities with NASH

A supervised PLS-DA was performed including all profiled lipids to examine if plasma OXLs profile can discriminate between ethnicities with NASH. The model demonstrated separation between HIS and CAU with 22 (49%) contributing lipids having VIP>1. This indicates differences in OXLs profile characterizes HIS and CAU with NASH (Figure S4 and Table S5). The Q^2^ and R^2^ for the model were 0.62 and 0.72, respectively, indicating a fair reliability of the model. Of note, an overlap between ethnic groups was observed and subjects within this area shared advanced fibrosis (grade 3 and 4), indicating that HIS and CAU subjects with NASH and advanced fibrosis share similar plasma OXLs profile. Also, it suggests that advanced fibrosis may be attenuating the multivariate model. Therefore, we repeated the analysis after excluding subjects with advance fibrosis (Figure 3 and Table S5). As a result, the model exhibited complete separation between ethnicities in NASH with 20 (44%) lipids contributing to this separation (VIP>1.0). The Q^2^, R^2^ were 0.99 and 0.98, respectively, indicating optimal prediction and reliability of the multivariate model.

**Figure 3.**
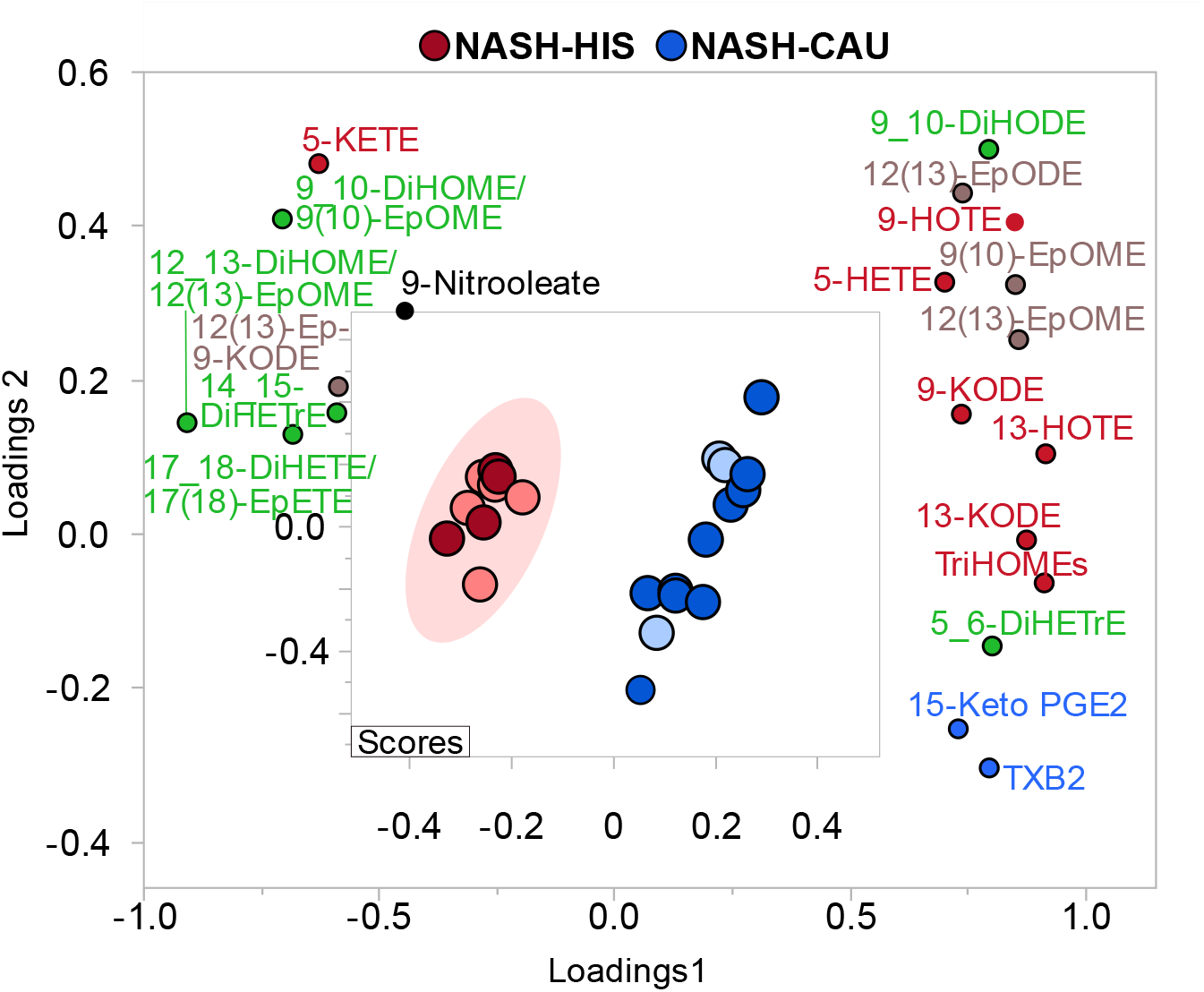
Supervised multivariate clustering model demonstrating ethnicity specific OXLs profile discriminates HIS and CAU with NASH without advanced fibrosis. Score plot for NASH-HIS (red) vs. NASH- CAU (blue) when excluding when excluding stage 3 and 4 fibrosis (*n* = 9 HIS and 13 CAU). Light color represents subjects from primary cohort and dark color represents secondary cohort. The model was performed on all profiled lipids. Lipid mediators are colored according to their metabolizing enzymes pathways, lipoxygenase, and autoxidation (red); cytochrome p450 epoxygenase (brown); and soluble epoxide hydrolase (green); cyclooxygenase (blue); other (black). The model was validated with leave-one-out cross validation (LOOCV). The (Q^2^) and (R^2^) is 0.99 and 0.98, respectively. Details on variable importance into the projection (VIP) scores and cross validation are shown in Table S6. Caucasian; HIS, Hispanics; NASH, Non-alcoholic steatohepatitis; DiHETE, dihydroxyeicosatetraenoic acid; DiHETrE, dihydroxyeicosatrienoic acid; DiHODE, dihydroxyoctadecadienoic acid; DiHOME, dihydroxyoctadecenoic acid; Ep-KODE, epoxyoxooctadecenoic acid; EpETE, epoxyeicosatetraenoic acid; EpODE, epoxyoctadecadienoic acid; EpOME, epoxyoctadecenoic acid; HETE, hydroxyeicosatetraenoic acid; HOTE, hydroxyoctadecatrienoic acid; KETE, keto-eicosatetraenoic; KODE, keto-octadecadienoic acid; PGE, prostaglandin E; TriHOME, trihydroxyoctadecaenoic acid; TXB, thromboxane.

In subjects with NASH and mild to moderate fibrosis, OXLs profiles showed opposite direction of change between ethnicities in some arachidonic acid-derived mediators, with TXB_2_, 15-Keto PGE_2_, 5-HETE and 5,6-DiHETrE being lower in HIS compared to CAU. Many OXLs derived via LOX pathway and/or auto-oxidative routes were lower in NASH-HIS, including TriHOMEs, 13-KODE, 9-KODE, 13-HOTE, 9-HOTE, and 5-HETE. Some CYP-derived OXLs were lower in NASH-HIS, as 9(10)-EpOME, 12(13)-EpOME and 12(13)-EpODE. Multiple sEH enzymatic indices were found higher in NASH-HIS, compared to NASH-CAU, including 12,13-DiHOME/12(13)-EpOME; 9,10-DiHOME/9(10)-EpOME; 17_18-DiHETE/17(18)-EpETE along with higher levels of the vicinal diol, 14,15-DiHETrE. These findings further confirm lower arachidonic acid OXLs in HIS with NASH, which are also characterized by lower LOX and higher sEH derived lipid mediators compared to CAU. They also indicate that plasma OXL profiles can discriminate between ethnicities in NASH.

## 4. Discussion

This study is the first to examine targeted plasma PUFAs, OXLs, and eCBs profiles with regards to ethnicity in a group of HIS and CAU subjects with obesity and biopsy-diagnosed NAFL and NASH. Our findings indicate that: 1) NAFL and NASH are characterized by ethnicity-related differences in plasma PUFA profiles, independent of histological severity; 2) Ethnicity-related differences in plasma OXLs profiles characterize NASH, independent of histological scores; 3) Plasma PUFA profile is altered in apparently healthy HIS, independent of obesity.

The hepatic and serum/plasma PUFA profiles are dysregulated in NAFLD [14–18]. Our results expand on these findings and show ethnicity-related differences in plasma PUFA profile in NAFL and NASH. With NAFL, both ethnicities showed higher α-linolenic and linoleic acid levels, which were pronounced in CAU but not in HIS. This can be attributed to differences in the levels of these PUFAs in lean and healthy subjects, with HIS having higher levels compared to their CAU counterpart. There was also a divergence in LC-PUFA profiles between ethnicities. CAU showed higher levels mainly affecting arachidonic acid, while HIS displayed lower levels mainly affecting docosahexaenoic and eicosapentanoic acids. Consistent trends were shown with the progression from 0-NASH to NASH with arachidonic almost reaching significance (*p-*Interaction =0.07). This is in line with our previous untargeted profiling in NAFL done on the same subjects showing higher C18-PUFAs in CAU, and a trend for lower LC-PUFA with tendencies shown for docosahexaenoic and eicosapentanoic acids in HIS [25]. The lack of significant difference in LC-PUFA in our previous analysis may be due to the semi-quantitative nature and the clustering statistical approach. Also in agreement with our current finding is the lower serum/plasma docosahexaenoic and eicosapentanoic levels reported in obese HIS, compared to non-HIS [34, 35]. Together, this implies diminished plasma LC-PUFA characterizes obese HIS with NAFL and NASH. As dietary intake affects PUFA circulating and tissue levels [36], the ethnicity-dependent differences in n-3 PUFAs intake which is reportedly lower in HIS could be responsible for these observed changes [35, 37]. While we did not account for diet, our findings suggest a possible role as we observed higher linoleic and α-linolenic acid levels independent of obesity in lean healthy HIS compared to CAU. Beside diet, genetic variants in cluster region of fatty acid desaturases *(FADS)* can predict LC-PUFA serum/blood levels [38, 39]. Single nucleotide polymorphisms (SNP) in *FADS1* and *FADS2*, *w*hich encode fatty acid desaturases, were robustly associated with NAFLD [40, 41]. Lower Δ-5 desaturase levels are reported in both NAFL and NASH [14, 15, 18]. Notably, SNPs in *FADS* that are associated with insufficient biosynthesis of LC-PUFA present with high frequency in Amerindians, a subgroup of HIS [42]. However, genotype was not examined in current study, and both ethnicities had lower estimated Δ-5 desaturase activity with NAFL. Therefore, diet and/or genetic factors may contribute to the observed ethnicity-related PUFA alterations but await further assessment.

Other key findings include the ethnicity-related differences in OXLs and eCBs profiles. The COX pathway exerts pro-inflammatory effects as it catalyzes the conversion of arachidonic acid to prostaglandin PGE_2_, thromboxane TXB_2_, and other fatty acid alcohols [8]. In animal models of NASH, the expression and activity of COX-2 were upregulated, and its inhibition ameliorated NAFL and NASH [43, 44]. Previously, high TXB_2_ and PGE_2_ levels were reported in subjects with NAFL and NASH [20]. Findings in NASH-CAU from our secondary analysis are consistent with this literature, as TXB_2_ and 15-Keto PGE_2_ discriminated between ethnicities with NASH with higher levels in CAU and lower levels in HIS. When comparing within ethnicities, the progression from 0-NASH to NASH in HIS was marked with a trend for lower arachidonic acid, almost reaching statistical significance (*p-*Interaction =0.07) and downstream OXLs with TXB_2_ being differentially lower (*p-*Interaction <0.05). These findings suggest ethnicity-related alteration in arachidonic acid metabolism and downstream COX-derived OXLs in NASH.

Evidence from animal studies indicates a role for LOX pathways in NAFL and inflammation [45, 46]. LOX pathways lead to the synthesis of fatty acid alcohols, ketones, hydroperoxides, and the specialized pro-resolving mediators (SPMs). With exceptions, n-6 PUFA derived alcohols are pro-inflammatory [8]. Under oxidative stress, PUFAs can also undergo auto oxidation to form alcohols, ketones, hydroperoxides [9]. Previous studies reported higher LOX and auto-oxidation metabolites in NAFL and increased arachidonic acid metabolites via LOX with the progression to NASH [16, 20,47]. In our results, compared to control groups, NAFL and NASH in both ethnicities presented higher alcohols and ketones derived from C18-PUFAs, indicating an upregulated LOX pathway(s). In NAFL, we observed a positive correlation between some fatty acid alcohols and the oxidative stress markers, F_2_-isoprostanes and 9-HETE (Table S6), implying a contribution of non-enzymatic auto-oxidation.

Interestingly, our secondary analysis showed many LOX derived OXLs being higher in NASH-CAU compared to NASH-HIS, with a similar trend found for the oxidative stress marker, 9-HETE (VIP=0.98). Together, these indicate that while LOX and oxidative pathways are upregulated with NAFL in both ethnicities, the magnitude of these alterations is lesser in HIS with NASH, compared to CAU. Based on this finding, we reasoned that LOX, and possibly oxidative stress, may be pivotal for NASH severity in CAU but not in HIS.

CYP enzymes catalyze the synthesis of fatty acids epoxides and alcohols. With some exceptions, fatty acid alcohols are pro-inflammatory and epoxides are anti-inflammatory, transient and rapidly hydrolyzed by the action of sEH to form inactive or less active vicinal diols [8, 48]. A role for sEH in NAFLD progression is indicated by animal studies, showing that sEH inhibition improves NAFL, NASH and fibrosis [48]. In subjects with NASH, compared to NAFL, arachidonic acid derived vicinal diols are higher [20]. Our results show, with the progression to NASH, an ethnicity-dependent opposite trend for vicinal diols derived from C18-PUFA, which were higher in HIS and lower in CAU. Some of these vicinal diols and sEH enzymatic indices showed interaction (ethnicity x NASH) and were found higher in NAHS-HIS and lower in NASH-CAU, compared corresponding 0-NASH. This may suggest higher activity of sEH in NASH-HIS. Our secondary analysis also shows higher ratios of multiple sEH enzymatic indices in HIS compared to CAU, and lower C18-PUFA epoxides possibly due higher hydrolysis rate. NASH-CAU showed higher levels of many PUFA epoxides, compared to NASH-HIS, indicating upregulated CYP pathway(s) and/or less hydrolysis. Together, our finding highlights ethnicity-related differences in sEH activity that was higher in HIS with NASH.

Extensive evidence from animal studies indicates a role for eCB system in NAFL, mitochondrial dysfunction and inflammation and fibrosis [49, 50]. In NAFL, both ethnicities had higher levels of several eCBs. However, many eCBs were higher in CAU and lower in HIS with the progression to NASH, compared to corresponding 0-NASH (raw *p-*Interaction <0.05). We also observed levels of the oleic acid-derived mediators N-oleoyl glycine, and oleoylethanolamide. These observations could not be examined in our secondary analysis as we detected limited numbers of eCBs and did not profile for fatty acids.

Nevertheless, this may indicate ethnicity-related variations in eCBs profiles and oleic acid metabolism with NAFLD in HIS that need to be further examined.

The observed ethnicity-related alterations may be relevant to NAFLD severity. Eicosapentaenoic and docosahexaenoic acids modulate hepatic fatty acid oxidation, *de novo* lipogenesis, redox balance and inflammation via direct interaction with nuclear receptors and transcription factors [51]. These LC-PUFAs are also precursors to potent SPMs which drive inflammatory resolution [8]. Also, the proinflammatory cascade of arachidonic acid via COX is necessary for the biosynthesis of SPMs and initiating inflammatory resolution [11, 52]. Therefore, a diminished level of these PUFAs may abolish anti-steatogenic and anti-inflammatory mechanisms. Likewise, a higher sEH activity may result in deactivation of anti-inflammatory PUFA epoxides [8, 48]. Interestingly, our findings suggest that upregulated LOX pathway(s) may be imperative to NASH severity in CAU with a lesser extent in HIS. Collectively, we postulate that the observed ethnicity-related changes translate to the more advanced NASH histological presentation seen in HIS. Of note, these changes are independent of fibrosis or NAS scores, in fact, histology adjustment resulted in stronger differences in both analyses, implying that subjects with advanced fibrosis may share similar lipidomic profile.

Our findings have clinical/diagnostic implications. The stage of liver fibrosis in NASH is critical to guide decisions on therapy [53]. Given that liver biopsy is subjected to risks and limitations, there is an ongoing search for noninvasive biomarker for NAFLD [54]. Several OXLs derived from arachidonic acid were shown to predict liver histological features. For example, NASH was characterized by a stepwise increase in 5- and 15-HETE; PGE_2_; and some vicinal diols [16, 20]. While our findings in NASH-CAU show trends consistent with current literature, findings in HIS indicate otherwise. Ethnicity-related differences in plasma metabolomic profile have been reported before in Alzheimer’s disease and bladder cancer [55, 56]. Based on our finding, we propose that ethnicity-specific plasma signature may characterize NASH. On another note, clinical evidence supports a role of eicosapentaenoic and docosahexaenoic acids supplementation in improving NAFL and its risk factors [57, 58]. Also, there is growing data on the utility of sEH inhibitors in NASH treatment with sEH inhibitors entering clinical trials [48, 59, 60]. Therefore, evaluating these interventions for the treatment of NASH, particularly in the HIS population seems warranted.

This “proof-of-concept” analysis is based on a small, single-center study. The limited sample size may have compromised the correction for multiple testing in the primary analysis. However, findings from the secondary analysis were consistent and the multivariate model is validated for overfitting and predictability. Other strengths include subjects were biopsy-characterized and analysis was adjusted for BMI and histology. In conclusion, we performed targeted lipidomic profiling for PUFAs and related lipid mediators with regards to ethnicity. Results show ethnicity-related divergence in LC-PUFA and downstream OXLs profiles with NAFL and NASH progression, independent of histological scores. Our secondary analysis indicates that in NASH and compared to CAU, HIS are characterized by lower levels of arachidonic acid derived OXLs, downregulated LOX with an upregulated sEH pathway(s). The implications of these differences on the disparity reported in NAFLD rate and severity are not clear but worth further investigations. Our findings suggest ethnicity-specific lipidomic signature may characterize NASH. Although preliminary, these novel observations support the need for larger validation studies.

## 5. Data Availability Statement

Analysis protocol and reported data are available at the Metabolomics Workbench (http://www.metabolomicsworkbench.org), study ID numbers (ST000977 and ST001845).

## 6. Conflict of interest

V.M. serves on the Advisory Board of Alexion Pharmaceuticals.

## 7. Author Contributions

Original draft writing, T.A.M.; Draft editing, T.A.M., V.M., K.B., J.W.N., Formal analysis and visualization, T.A.M., K.B; Methodology, T.A.M, V.M., O.F., D.K, K.M and Y.Y.W.; Investigation, T.A.M, K.M.; Resources, V.M., O.F., S.S., K.L.S., P.J.H. and M.A.; Funding Acquisition-V.M., J.W.N. and P.J.H.

## 8. Funding

This work was funded by The National Institutes of Health (NIH)-West Coast Metabolomic Center (WCMC) grant number U24 DK097154, by R01 DK104770 (to V.M.), by 1R01 HL09133 (to P.H.) and 1R01 HL107256 (to P.H.). Additional fund was provided by The United States Department of Agriculture (USDA) project 2032-51530-025-00D (to J.W.N). The USDA is an equal opportunity provider and employer. The content is exclusively the responsibility of the authors and does not represent the official views of the NIH, the WCMC or the USDA.

## Abbreviations

0-NASH: NASH-free
CAU: Caucasian
COX: Cyclooxygenases
CYP: Cytochrome P450
eCBs: Endocannabinoids
FADS: Fatty acid desaturases
FC: Fold change
FDR: False discovery rate
HC: Healthy control
HIS: Hispanic
LOOCV: leave- one-out cross validation
LOX: Lipoxygenases
NAFL: Steatosis
NAFLD: Non-alcoholic fatty liver disease
NAS: The NAFLD Activity Score
NASH: Non- alcoholic steatohepatitis
n-3 PUFA: Omega−3 polyunsaturated fatty acids
n-6 PUFA: Omega−6 polyunsaturated fatty acids
NASH-CRN: The NASH Clinical Research Network
OXLs: Oxylipins
PCA: Principal component analysis
PLS-DA: Partial least square-discriminant analysis
PUFA: Polyunsaturated fatty acids
sEH: Soluble epoxide hydrolases
SPMs: Specialized pro-resolving mediators
SNP: single nucleotide polymorphisms
VIP: Variable importance in projection
Δ-5 desaturase: Delta fatty acid desaturase-5.

## Supplemental Figure

**Figure S1.**
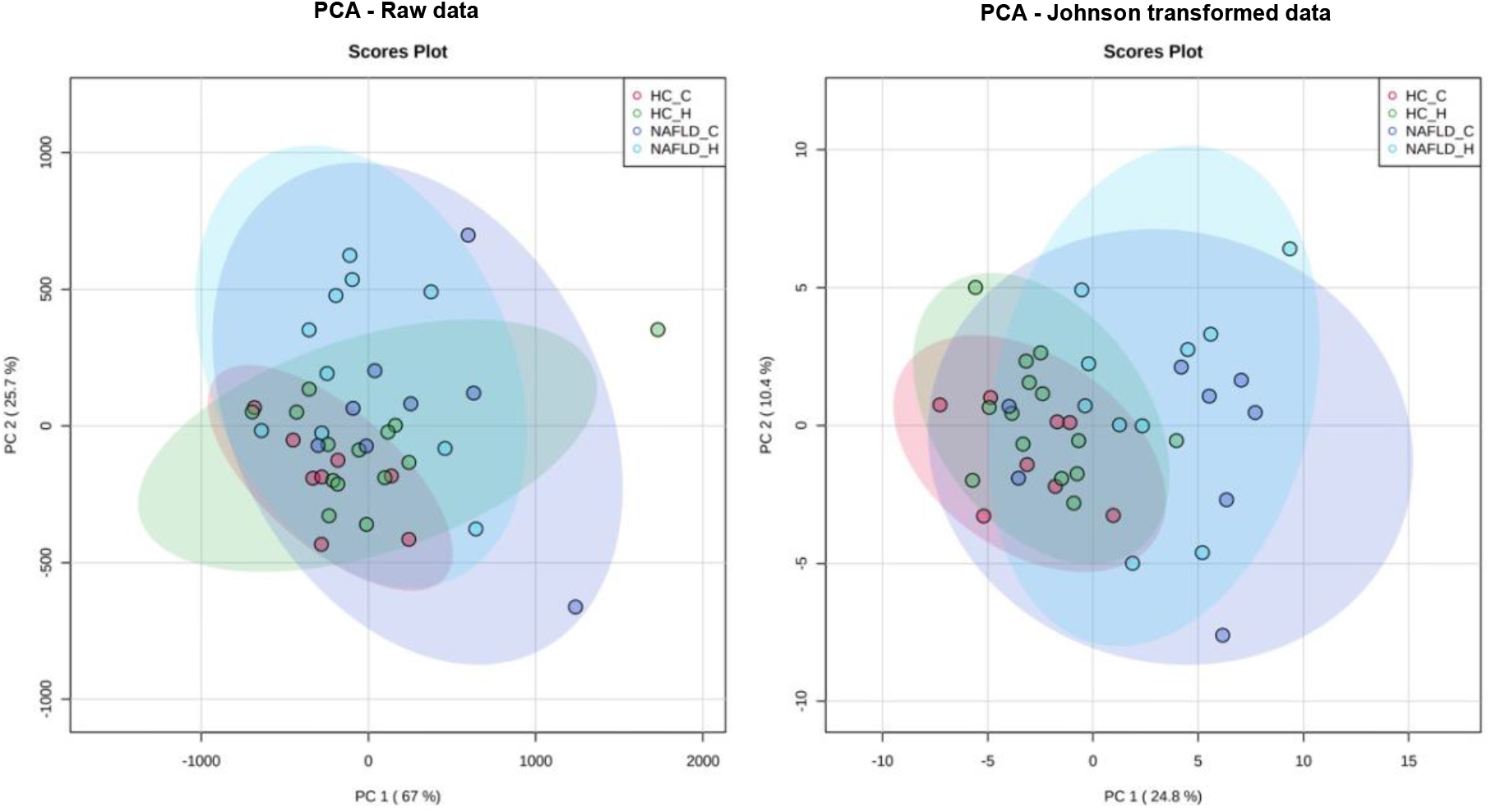
Principle component analysis (PCA) illustrating outliers before (left) and after (right) data normalization.

**Figure S2.**
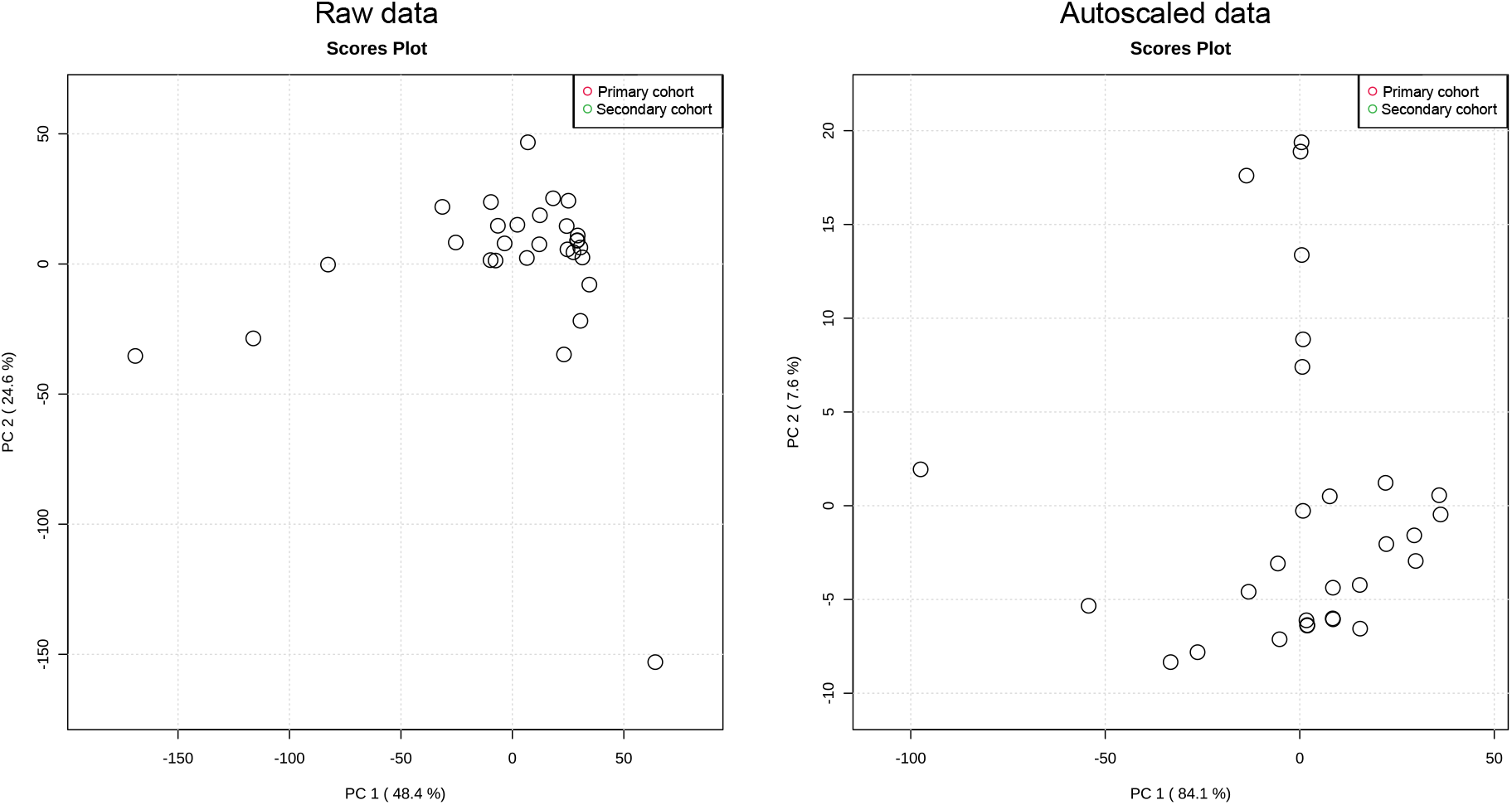
Principle component analysis (PCA) illustrating the unsupervised clustering of samples from primary (red) and secondary cohort (green) shown before (left) and after (right) batch correction.

**Figure S3.**
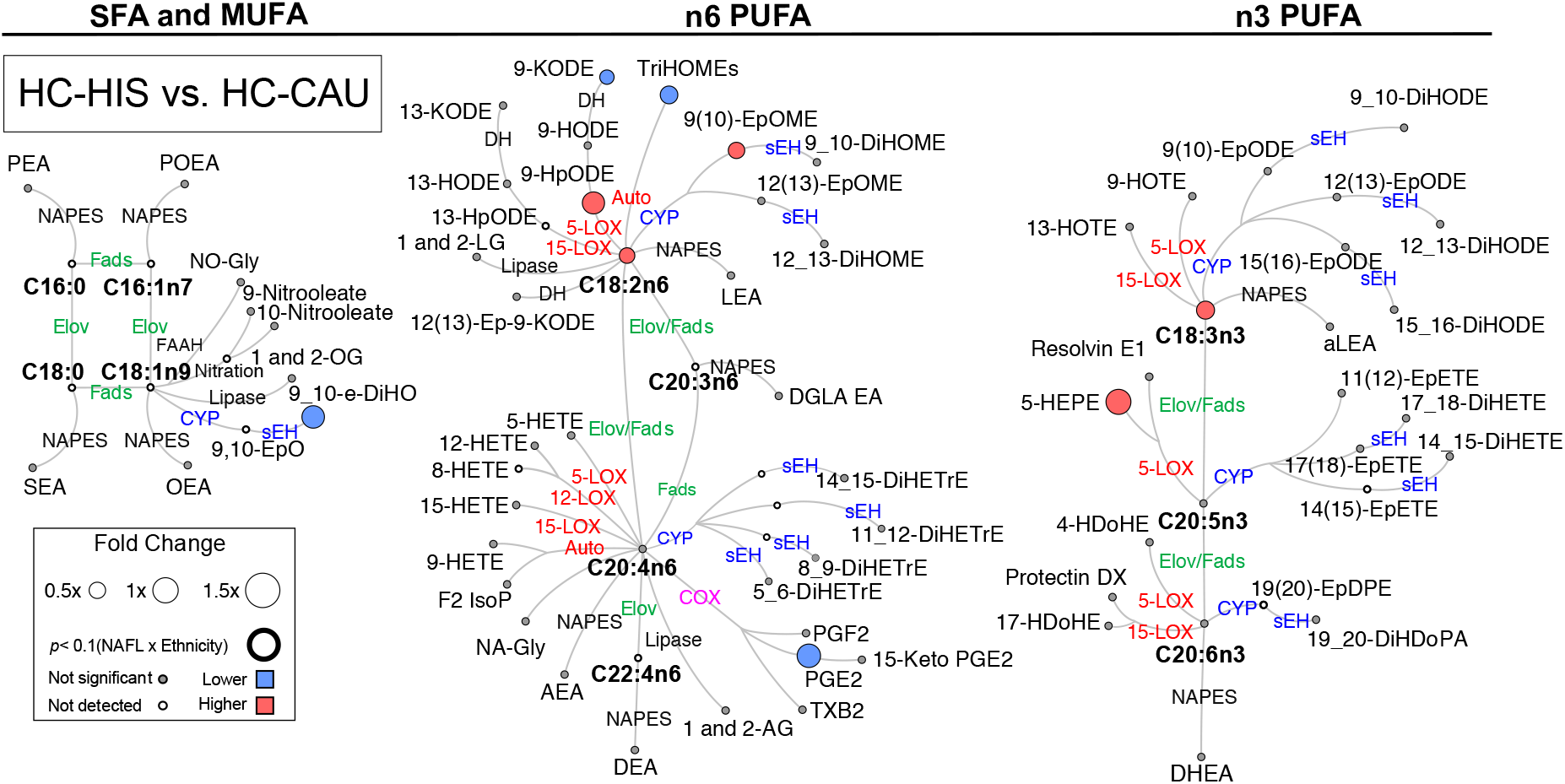
Differences in plasma PUFA and lipid mediators between HIS and CAU in lean healthy subjects. Metabolic network illustrating saturated (SFAs), monounsaturated (MUFAs) and polyunsaturated fatty acids (PUFAs), including n-3 and n-6 fatty acids with pathway of oxylipins and endocannabinoids synthesis. Node size represents the fold change of lipids, calculated for each ethnicity separately as (HIS - CAU)/CAU. Node’s color represents the directionality of metabolite differences: higher in NAFL (red); lower in NAFL (blue); no change (grey). Only fold change for lipids with differences between HIS vs. CAU (*t*-test raw *p* <0.05) are shown. Lipids means and corresponding *p*-values are detailed in Table S3. (*n* =14 HIS and 8 CAU). Fatty acids are described by number of carbons and double bounds of the fatty acyl moiety (i.e., C18:2n6). CAU, Caucasian; HC, Healthy control; HIS, Hispanics; ADH, alcohol dehydrogenase; AEA, arachidonoyl ethanolamine; AG, arachidonoyl glycerol; Auto, autoxidation; COX, cyclooxygenase; CYP, cytochrome P450; DEA, docosatetraenyl ethanolamide; DGLEA, dihomo-gamma-linolenoyl ethanolamide; DH, dehydrogenase; DHEA, docosahexaenoyl ethanolamide; DiHDoPA, dihydroxydocosapentaenoic acid; DiHETE, dihydroxyeicosatetraenoic acid; DiHETrE, dihydroxyeicosatrienoic acid; DiHO, dihydroxyoctadecanoic acid; DiHODE, dihydroxyoctadecadienoic acid; DiHOME, dihydroxyoctadecenoic acid; Elov, fatty acid elongase; Ep-KODE, epoxyoxooctadecenoic acid; EpETE, epoxyeicosatetraenoic acid; EpODE, epoxyoctadecadienoic acid; EpOME, epoxyoctadecenoic acid; F2-IsoP, F2 isoprostanes; FAAH, fatty acid amide hydrolase; Fads, fattyaciddesaturase; HDoHE, hydroxydocosahexaenoic acid; HEPE, hydroxyeicosapentaenoic acid; HETE, hydroxyeicosatetraenoic acid; HODE, hydroxyoctadecadienoic acid; HOTE, hydroxyoctadecatrienoic acid; HpODE, hydroperoxyoctadecadienoic acid; KETE, keto-eicosatetraenoic; KODE, keto-octadecadienoic acid; LEA, Linoleyl ethanolamine; LG, linoleoylglycerol; LOX, lipoxygenase; MUFA, Monounsaturated fatty acid; NA-Gly, arachidonylglycine; NAPES, N-acylphosphatidyl ethanolamine-specific; NO-Gly; OEA, oleoyl ethanolamine; OG, oleoylglycerol; PEA, palmitoyl ethanolamine; PGE, prostaglandin E; PGF, prostaglandin F; POEA, palmitoleoyl ethanolamide; PUFA, Polyunsaturated fatty acids; SEA, stearoyl ethanolamide; sEH, soluble epoxide hydrolase; SFA, Saturated fatty acids; TriHOME, trihydroxyoctadecaenoic acid; TXB, thromboxane.

**Figure S4.**
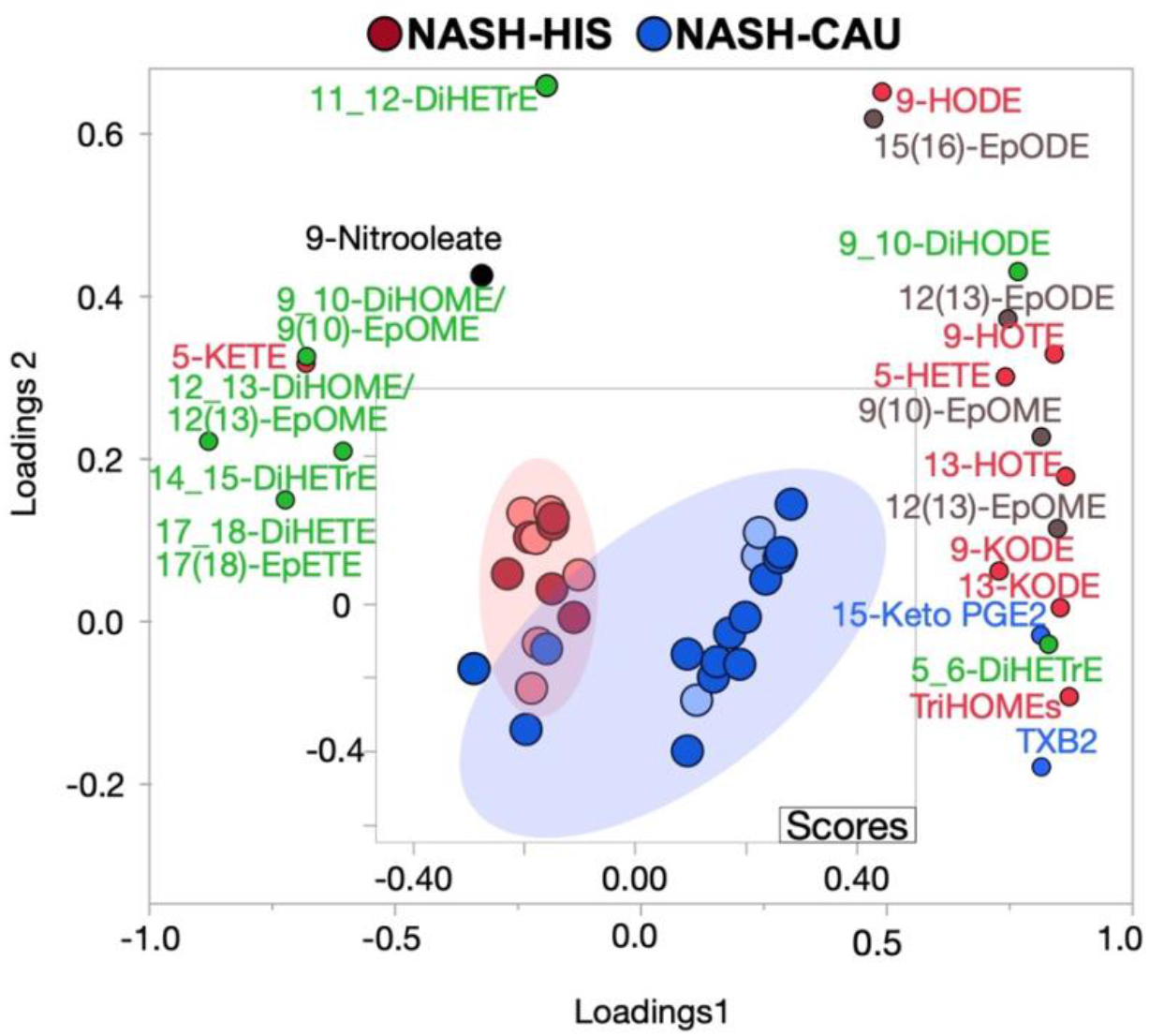
Supervised multivariate clustering model demonstrating comparing OXLs profile in HIS and CAU with NASH and advance fibrosis. Score plot for NASH-HIS vs. NASH-CAU when including stage 3 and 4 (*n* = 12 HIS and 17 CAU). Light color represents subjects from primary cohort and dark color represents secondary cohort. The model was performed on all profiled lipids. Lipid mediators are colored according to their metabolizing enzymes pathways, lipoxygenase, and autoxidation (red); cytochrome p450 epoxygenase (brown); and soluble epoxide hydrolase (green); cyclooxygenase (blue); other (black). The model was validated with leave-one-out cross validation (LOOCV). The (Q^2^) and (R^2^) is 0.62 and 0.72. Details on variable importance into the projection (VIP) scores and cross validation are shown in Table S6. CAU, Caucasian; HIS, Hispanics; DiHETE, dihydroxyeicosatetraenoic acid; DiHETrE, dihydroxyeicosatrienoic acid; DiHODE, dihydroxyoctadecadienoic acid; DiHOME, dihydroxyoctadecenoic acid; EpETE, epoxyeicosatetraenoic acid; EpETrE, epoxyeicosatrienoic acid; EpODE, epoxyoctadecadienoic acid; EpOME, epoxyoctadecenoic acid; HETE, hydroxyeicosatetraenoic acid; HODE, hydroxyoctadecadienoic acid; HOTE, hydroxyoctadecatrienoic acid; KETE, keto-eicosatetraenoic; KODE, keto-octadecadienoic acid; PGE, prostaglandin E; TriHOME, trihydroxyoctadecaenoic acid; TXB, thromboxane.

## Supplemental tables captions

**Table S1. Details on detected lipid class and identifiers.**

**Table S2. Targeted quantification data table for comparisons of Hispanic (HIS) and Caucasian (CAU) obese subjects with Non-alcoholic fatty liver disease (NAFL) vs. lean healthy control (HC).** Comparisons between NAFL vs. HC performed separately in each ethnicity. (*n* =10 HIS and 8 CAU); HC (*n* =14 HIS and 8 CAU). Details are shown for *p*-values; means (upper-lower 95% mean); fold change (FC); *t*-test and ANCOVA *p*-values for individual lipids. Values are rounded to 3 significant figures.

**Table S3. Demographic, clinical, and histological characteristics of histology-matched subjects.** General characteristics of NAFL and HC groups in both ethnicities shown as percent (for categorical data) and mean ± SEM (for nominal data). Comparisons were performed by *t*-test (nominal) or chi-square test (categorical). ALT - alanine aminotransferase; AST - aspartate aminotransferase; BMI - body mass index; CAU - Caucasian; DM -diabetes mellitus; FBG - fasting blood glucose; HbA1c - hemoglobin A1c; HDL - high-density lipoprotein, HIS -Hispanic; LDL - low density lipoprotein; NAFL – Non-alcoholic fatty liver; NAS - the NAFLD Activity Score; TG, triglycerides. *^(a)^* NAFL-HIS vs. HC-HIS; *^(b)^* NAFL-CAU vs. HC-CAU; *^(c)^* NAFL-HIS vs. NAFL-CAU.

**Table S4. Targeted quantification data table for comparisons of Hispanic (HIS) and Caucasian (CAU) with Non-alcoholic steatohepatitis (NASH) vs. 0-NASH.** Comparisons between NASH vs. 0-NASH performed separately in each ethnicity. NASH (*n* =6 HIS and 3 CAU); 0-NASH (*n* =4 HIS and 5 CAU). Details are shown for *p*-values; means (upper-lower 95% mean); fold change (FC); *t*-test and ANCOVA *p*-values for individual lipids. Values are rounded to 3 significant figures.

**Table S5: Targeted quantification data table for comparisons of Hispanic (HIS) and Caucasian (CAU) with Non-alcoholic steatohepatitis (NASH) in the secondary analysis.** Details on variable importance into the projection (VIP) scores and cross validation for multivariate clustering model demonstrating ethnicity specific OXLs profile discriminates HIS and CAU with NASH. *^(a)^* including stage 3 and 4 (*n* = 12 HIS and 17 CAU); *^(b)^* when excluding when excluding stage 3 and 4 (*n* = 9 HIS and 13 CAU). CAU - Caucasian; HIS - White Hispanic; Non-alcoholic steatohepatitis.

**Table S6: Pearson’s correlation analysis between LOX metabolites and markers of oxidative stress.** Correlation analysis between alcohols and ketones derived from 5, 12 and 15 LOX with and autoxidative markers, F2 isoprostanes and 9-HETE in *^(a)^* NAFL subjects (examined in primary cohort); *^(b)^* NASH subjects (examined in secondary cohort). CAU - Caucasian; HIS - White Hispanic; NAFL – Non-alcoholic fatty liver; Non-alcoholic steatohepatitis.

## Notes

### Author Declarations

All subjects were consented and the Institutional Review Board at the University of California, Davis approved the study protocol (# 856052).

